# Post-Traumatic Stress Disorder in Formerly Homeless Queer Students Enrolled at sub-Saharan African Higher Education Institutions: A Systematic Review Protocol

**DOI:** 10.1101/2025.09.16.25335905

**Authors:** Senzelokuhle. M. Nkabini, Lindiwe Nkabini-Anderson, Fezile Zuma

**Affiliations:** Faculty of Humanities, University of Pretoria. Hatfield Campus: Hatfield, Lynnwood Rd & Roper St, Pretoria, 0002. Gauteng province, South Africa; Clean Up Project,728 P Imfume Main Road, Amanzimtoti,4125 KwaZulu-Natal: South Africa

**Keywords:** Post-Traumatic Stress Disorder, PTSD, Formerly Homeless, Queer Students, Sub-Saharan Africa, Higher Education Institutions

## Abstract

**Background:** Post-traumatic stress disorder (PTSD) has continuously been an overlooked public health issue in sub-Saharan Africa (SSA), despite the documented effects this mental disorder has had on the 1.6 billion people that occupy this region. Globally, 80% of the world’s population, in low-and-middle-income countries, live with an untreated mental health issue. SSA has approximately 41 countries that are defined as low income (LIC’s) and lower-middle income (LMIC’s). 83% of (LIC’s) and 78% of (LMIC’s) have a national mental health policy and implementation strategy. However, only 3% of LIC’s and 14% of LMIC’s have successfully implemented these policies and guiding principles. In addition, LIC’s only have 0,8% mental health workers, while LMIC’s have reported of only retaining 2,3% workers that specialise in mental healthcare. This is unfortunate because 90% of SSA countries have made efforts to eliminate user fees at healthcare facilities. The ineffective execution of mental health policy plans is catastrophic for people that live in SSA, especially for socioeconomically marginalised groups such as homeless people, and queer individuals. These two groups have a higher rate of PTSD because of the numerous difficulties they encounter whilst trying to access healthcare facilities in this region. 70,3% of countries in Europe and 73% of countries in North America offer queer individuals protection against stigma and discrimination such as queerphobia in educational and social settings, combined with the criminalisation of this form of discrimination. On the other hand, in SSA queer students that have previously experienced homelessness, and are now enrolled at an HEI encounter political (besmirching state policies), economic (limited access to resources due to structural queerphobia), societal (discrimination and social exclusion) and health implications (continuous rise in the student’s mental health issues).

**Methods:** The primary aim of this systematic review is to map out and synthesise evidence through conducting a multi-database investigation for both qualitative, and quantitative studies on post-traumatic stress disorder in formerly homeless queer students enrolled at sub-Saharan African higher education institutions. The following databases will be utilized to search for studies: PubMed, PsycINFO, ProQuest, ERIC (Education Resources Information Center), Cochrane Reviews, WHO, UNHCR, UNFPA, UNAIDS, UNESCO, ACHPR, UN-Habitat, Sociology Database and Scopus. The Preferred Reporting Items for Systematic and Meta Analyses (PRISMA) ScR flow chart/diagram presented in figure 1 will be utilized to summarize the study selection process.

**Conclusion:** The proposed systematic review will generate findings pertaining to PTSD, and describe how this mental disorder affects formerly homeless queer students enrolled at SSA HEI’s. these findings will/can reveal the current existing literature gaps, regarding PTSD as a whole, and also the lack in research/studies focusing on formerly homeless queer students enrolled at HEI’s. Through addressing these gaps in this systematic review, humanitarian organisations can conduct further research on this topic. Moreover, through further attention being afforded to this topic, numerous amendments can be made with regards to WHO’s *Comprehensive Mental Health Action Plan 2013-2030* and the SDG’s 3,4,5,10 and 11.

**Systematic reviews registration:** PROSPERO CRD420251141977

## 1. Background

Post-traumatic stress disorder (PTSD) has continuously been an overlooked public health issue in sub-Saharan Africa (SSA), despite the documented effects this mental disorder has had on the 1.6 billion people that occupy this region. Globally, 80% of the world’s population, in low-and-middle-income countries, live with an untreated mental health issue (1,2). SSA has approximately 41 countries that are defined as low income (LIC’s) and lower-middle income (LMIC’s) (1,2). 83% of (LIC’s) and 78% of (LMIC’s) have a national mental health policy and implementation strategy (2,31). However, only 3% of LIC’s and 14% of LMIC’s have successfully implemented these policies and guiding principles (2,31). In addition, LIC’s only have 0,8% mental health workers, while LMIC’s have reported of only retaining 2,3% workers that specialise in mental healthcare (2,31). This is unfortunate because 90% of SSA countries have made efforts to eliminate user fees at healthcare facilities (3). The ineffective execution of mental health policy plans is catastrophic for people that live in SSA, especially for socioeconomically marginalised groups such as homeless people, and queer individuals (4). These two groups have a higher rate of PTSD because of the numerous difficulties they encounter whilst trying to access healthcare facilities in this region (4). In SSA these difficulties are prompted by the stigma, discrimination, and social exclusion rendered towards individuals that are homeless and/or identifying as queer (5). Moreover, this form of condemnation in some SSA countries, is state-sponsored by political leaders and reigning governments. This has led to both a denial and violation of human rights for homeless and queer identifying individuals trying to seek mental healthcare. Regrettably, this form of persecution continues to exist even for formerly homeless queer students enrolled in SSA higher education institutions (HEI’s).

Within a vast majority of upper-middle and developed countries, formerly homeless queer students enrolled in HEI’s encounter numerous challenges that affect their academic performance, and social skills due to the shame and stigma associated with being homeless in the past (6). However, 70,3% of countries in Europe and 73% of countries in North America offer queer individuals protection against stigma and discrimination such as queerphobia in educational and social settings, combined with the criminalisation of this form of discrimination (7). On the other hand, in SSA queer students that have previously experienced homelessness, and are now enrolled at an HEI encounter political (besmirching state policies), economic (limited access to resources due to structural queerphobia), societal (discrimination and social exclusion) and health implications (continuous rise in the student’s mental health issues) (7,30). Additionally, only one country (South Africa) in SSA offers queer individuals constitutional protection against discrimination, prohibits the incitement of violence against queer people, and has a criminal liability law on the basis of the victim’s sexual orientation or gender identity (7). Predictably, out of the 48 SSA countries (South Africa excluded) only 6% have a criminal liability law on the basis of violence perpetrated against queer victims, and only 4% within this region prohibit the incitement of violence or discrimination towards individuals that do not ascribe to heteronormative gender or sexual identities (8). Moreover, only three countries in SSA offer queer individuals protection against discrimination in housing/ accommodation/ shelter (8). Thus, the current lack in continental (Africa), regional (SSA) and state (SSA country) legislations that address this form of systemic inequity, and how it eventually leads to PTSD in formerly homeless queer students enrolled at SSA HEI’s.

The *Comprehensive Mental Health Action Plan 2013-2030* program was introduced and implemented by the World Health Organisation (WHO), in order to increase availability & user-friendliness of mental healthcare facilities, reduce the stigma associated with mental disorders, and the acknowledgement of patients’ human rights (9). In the case of formerly homeless queer students enrolled at SSA HEI’s, this program can aid or is currently assisting with improving college/university centres for student counselling and career development. However, within the SSA region, formerly homeless queer students enrolled at HEI’s still continue to encounter numerous forms of queerphobia, stigma and violent physical discrimination (10). This form of aggression is commonly administered by fellow students, lecturers, graduate studies supervisors, faculty deans, departmental heads, college/university administration staff, college/university security personnel, and other individuals stationed in different college/university departments (11,26,27,28,29). The existence of queerphobia, stigma and violent physical discrimination in SSA HEI’s is expected, unlike their international counterparts where this is a highly contested subject. This is due to HEI’s being viewed as beacons of freedom of speech, coupled with their national and international mandates of recognizing, valuing and being inclusive of all enrolled students (11,26,27,28,29). This highly contested subject is considered a global challenge by the United Nations (UN), and is included in some of the 17 sustainable development goals (SDGs) that were created to address the numerous social, economic, and health-related issues that are also experienced by people living in the SSA region (12). PTSD affecting formerly homeless queer students enrolled in SSA HEI’s is both a global and regional challenge. Hence, it is included in SDGs such as SDG 3: *Good Health and Well-being*, SDG 4: *Quality Education*, SDG 5: *Gender Equality*, SDG 10: *Reduced Inequality*, and SDG 11: *Sustainable Cities and Communities*. These SDGs are relevant because they promote the provision, and access to quality healthcare & education, as well as equality & non-discrimination of any individual or group. They also ensure that continental, regional and state humanitarian organisations create communities that are sustainable, safe and inclusive.

In order to systematically map and synthesize evidence of PTSD in formerly homeless queer students enrolled at SSA HEI’s, and identify literature gaps that could inform future research the primary research question guiding this review is: What evidence exists regarding PTSD in formerly homeless queer students enrolled at SSA HEI’s? The secondary research question is: What evidence exists regarding the factors that contribute to PTSD in formerly homeless queer students enrolled at SSA HEI’s? The duplication of previous reviews and studies has been avoided through conducting a preliminary database search, through PubMed, Cochrane Reviews database of systematic reviews, and several other global databases. According to the research team (SMN, LN-A & FZ), a systematic review pertaining to mental health has been conducted on queer identities in sub-Saharan Africa for a study titled *A systematic review of university students mental health in sub-Saharan Africa* (13). However, based on the research teams’ knowledge, there has been no systematic review that has been conducted regarding PTSD in formerly homeless queer students enrolled at SSA HEI’s. Hence, the importance and relevance of this review. Moreover, the results from this systematic review will inform upcoming research that is conducted by global, continental (Africa), and regional (SSA) humanitarian organisations such as WHO, the Joint United Nations Programme on HIV/AIDS (UNAIDS), the United Nations High Commissioner for Refugees (UNHCR), the United Nations Population Fund (UNFPA), the United Nations Educational, Scientific and Cultural Organization (UNESCO), the United States Presidents Emergency Plan for Aids Relief (PEPFAR), the African Commission on Human and People’s Rights (ACHPR), United Nations Human Settlements Programme (UN-Habitat), and International Lesbian, Gay, Bisexual, Trans and Intersex Association (ILGA).

## 2. Methods

The primary aim of this systematic review is to map out and synthesise evidence of, PTSD in formerly homeless queer students enrolled at SSA HEI’s from existing literature. All forms of studies, grey literature and peer-reviewed journal articles focusing on PTSD, and formerly homeless queer students enrolled at SSA HEI’s will be sourced. The primary research question that will guide this review is: What evidence exists regarding PTSD in formerly homeless queer students enrolled at SSA HEI’s? The secondary research question is: What evidence exists regarding the factors that contribute to PTSD in formerly homeless queer students enrolled at SSA HEI’s? The eligibility of the research question was adequately addressed by the P.I.Co (population, interest, context) framework. This framework was selected because it has been adapted from the P.I.C.O framework, which is used for quantitative systematic reviews (14). Thus, In order to accommodate qualitative systematic reviews a P.I.Co framework was created (14).

### P.I.Co (Population, Interest, Context) framework

#### Population

Formerly homeless queer students that are either same-sex attracted, non-binary, transgender, intersex or asexual (15).

Age: 18 to 80+ years old is the average age range that will be used to filter the literature from the databases, because most students are enrolled at this age in HEI’s after graduating from high school (16).

#### Interest

PTSD is a mental disorder that develops after an individual experiences a traumatic or life-threatening past event, that consistently replays in their present life either through flashbacks, nightmares, or when they are partaking in certain activities that link to the traumatic event (2,4).

#### Context

SSA HEI’s established within the 49 countries that are located in the African continent (17).

#### Sources of evidence

All studies, grey and empirical literature containing evidence on PTSD in formerly homeless queer students enrolled at SSA HEI’s.

**Publication Year Range:** 01/01/2016-07/09/2025 **Language:** No language restriction

### 2.1 Information sources and search strategy

All searches will be conducted by SMN, LN-A and FZ for studies and study designs published in peer-reviewed journals, grey literature, published and unpublished dissertations, case studies, reviews, essays, theses and symposium abstracts. The following databases will be utilized to search for studies: PubMed, PsycINFO, ProQuest, ERIC (Education Resources Information Center), Cochrane Reviews, WHO, UNHCR, UNFPA, UNAIDS, UNESCO, ACHPR, UN-Habitat, Sociology Database and Scopus. The search strategy will use the Boolean term ‘OR’ to separate words or search terms. The following search terms will be utilised: post-traumatic stress disorder/PTSD OR formerly homeless OR queer OR students OR sub-Saharan African/SSA OR higher education institutions/HEI’s. A preliminary data base search was conducted by the research team using the search terms, and the results are presented in Table 1. The preliminary search produced all studies, qualitative and quantitative full-text grey literature and peer-reviewed literature with no language restrictions, and published within the search time-line from 01January 2016 till 07 September 2025.

**Table 1:** Preliminary database search results.

| Search terms | Date of Search | Databases | No. Retrieved |
| --- | --- | --- | --- |
|  | 01/01/2016 |  |  |
| post-traumatic stress | 07/09/2025 | PubMed | 2009 |
| disorder OR PTSD |  | Cochrane Reviews | 90 |
| OR formerly homeless OR queer |  | WHO | 10 |
| OR students OR sub-Saharan Africa |  |  |  |
| OR SSA OR higher education institutions |  |  |  |
| OR HEI's |  |  | Total: 2109 |

**Table 2:** Data charting table.

|  |
| --- |
| Author and year of publication |
| Article title |
| Study aim |
| Country |
| Study design/methodology |
| Study setting |
| Study population |
| Duration of the study |
| Factors contributing to PTSD |
| Population age |
| Relevant/significant findings |
| Key conclusions of article |
| Notes |

### 2.2 Inclusion and exclusion criteria

The selection of eligible studies will be based on the title (post-traumatic stress disorder/PTSD), abstract, setting (SSA/sub-Saharan African HEI’s/higher education institutions), study population (formerly homeless queer students) and the findings in full-text studies, books, grey literature and journal articles relating to PTSD, and formerly homeless queer students enrolled at SSA HEI’s. All database searches will be conducted on a weekly basis by SMN, LN-A and FZ to ensure new literature is included. Only studies that adhere to the following criteria will be included: (i) PTSD, and formerly homeless queer students enrolled at SSA HEI’s, (ii) qualitative and quantitative study designs iii) studies and articles published from 2016 to 2025 and, (iv) only studies, books, grey literature or journal articles that are restricted to human ages 18 to 80+ years old, full-text, have references/citations and have been peer-reviewed. Studies that are: (i) published before 2016, will be excluded.

### 2.3 Data management and study selection

The framework, enhancements to the framework and guidelines provided by Arksey and O’Malley’s (18), Levac *et al* (19), Daudt *et al* (20), and Johanna Briggs (21) will guide this review. The Preferred Reporting Items for Systematic and Meta Analyses (PRISMA) ScR flow chart/diagram presented in figure 1 will be utilized to summarize the study selection process (22,23). The authors and the two research assistants will ensure that all retrieved literature will be exported and saved to an Endnote 21.3 library folder, in order to enable the study to be reproduced for a second time at a later stage. Moreover, this process will allow SMN, LN-A and FZ to create separate libraries for each database, in order to import references, remove duplicates, and organize the findings.

**Figure 1:**
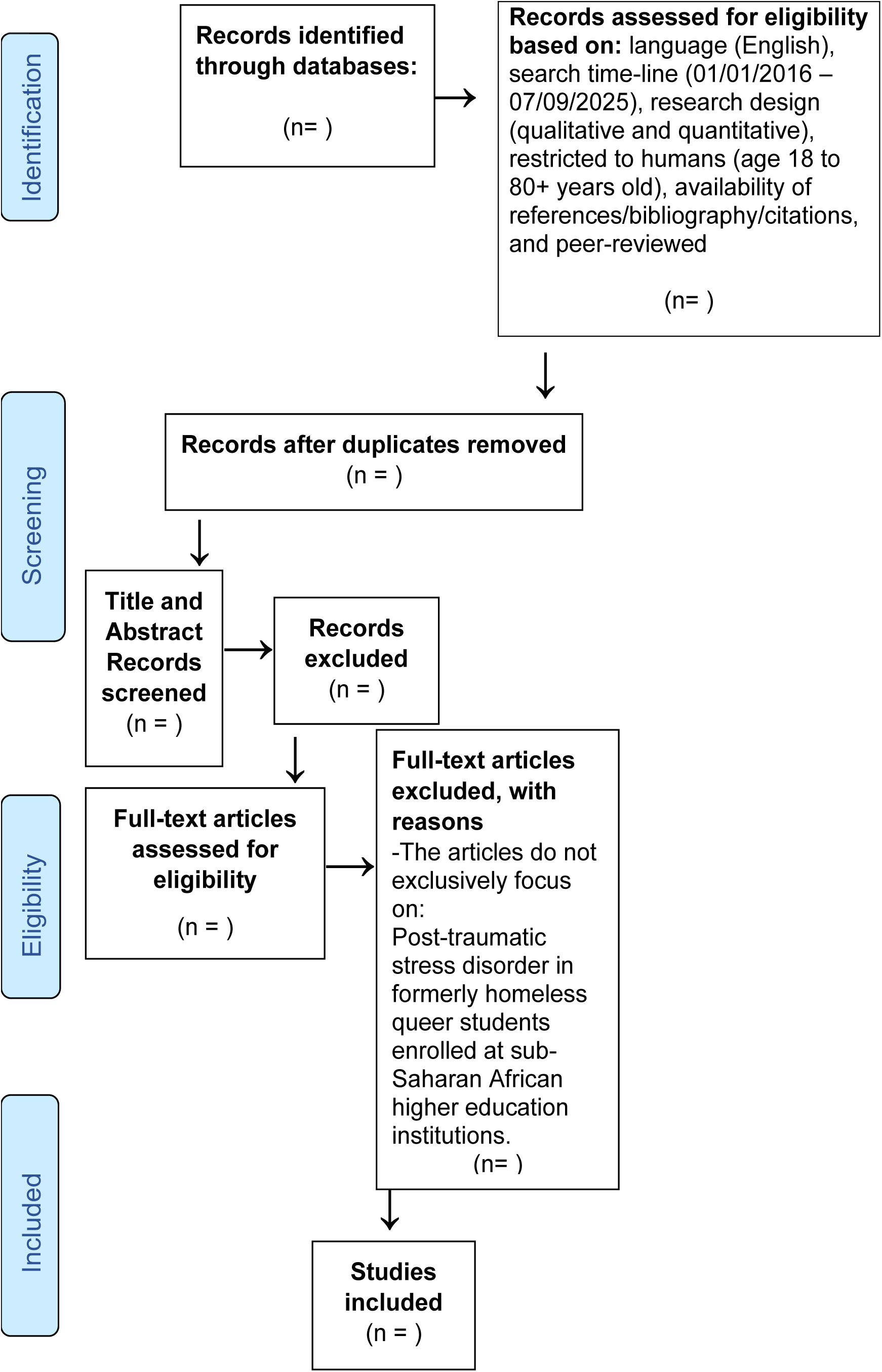
The Preferred Reporting Items for Systematic and Meta Analyses (PRISMA) ScR flow chart/diagram.

### 2.4 Data extraction

A data charting table will be used in order to document the extracted data. Studies that meet the inclusion criteria will be included on the charting form. The use of the NVivo data analysis software and Braun & Clarke’s (24) thematic framework will guide the qualitative analysis. The data extraction form will be updated by SMN, LN-A and FZ on a weekly basis to guarantee accurateness.

### 2.5 Risk of bias assessment

Studies or articles that only feature few countries that are located in the SSA region will not be included, because the research results from these studies do not represent SSA as a whole. In order to avoid biasness the review will utilise the mixed-method appraisal tool (MMAT) version 2018 to appraise the quality of all included evidence (25). SMN, LN-A & FZ will be responsible for assigning ratings of 100% for high average sources, 50% average and 25% for low-quality articles.

### 2.6 Discrepancies between the protocol and the systematic review

Discrepancies between the protocol, the actual review and the reasons and consequences thereof will be reported in the final report.

## 3. Data synthesis

A narrative synthesis will be conducted, which will provide texts and tables in order to synthesize and discuss the data of the studies and the methods, as previously described in the data extraction section.

## 4. Results

The results of this systematic review will be disseminated through publication in peer-reviewed scientific journals. The data will also be made available to humanitarian organisations such as WHO, UNAIDS, UNESCO, UNFPA, UNHCR, ACHPR, UN-Habitat and ILGA. The findings from this study will inform the future research projects that these organisations embark on.

## 5. Discussion

The proposed systematic review will generate findings pertaining to PTSD, and describe how this mental disorder affects formerly homeless queer students enrolled at SSA HEI’s. These findings will/can reveal the current existing literature gaps, regarding PTSD as a whole, and also the lack in research/studies focusing on formerly homeless queer students enrolled at HEI’s. Through addressing these gaps in this systematic review, humanitarian organisations such as WHO, UNESCO, UNAIDS, UNFPA, UNHCR, PEPFAR. ACHPR, ILGA and UN-Habitat can conduct further research on this topic. Moreover, through further attention being afforded to this topic, numerous amendments can be made with regards to WHO’s *Comprehensive Mental Health Action Plan 2013-2030* and the SDG’s 3,4,5,10 and 11. Furthermore, the results of this study will be disseminated through publication in peer-reviewed scientific journals, and the numerous United Nations humanitarian organisations that work at a continental level (Africa), regional level (SSA), and state level (country). These organisations have easier access to the affected populations, because they have representatives working within the numerous cities, suburban neighbourhoods, and rural communities. With regards to this study, organisations such as WHO, UNAIDS, UNESCO, and ILGA would utilize the results from this systematic review to assist non-governmental organisations (NGO’s), and non-profit organisations (NPO’s), as well as HEI’s student counselling departments, student leadership bodies, and student community organisations. Government departments within the various countries located in the SSA region, can also utilize these results to address the exacerbating homelessness issue in their metropolitan cities.

## Data Availability

All data produced in the present work are contained in the manuscript

## Acknowledgments

The authors express their gratitude to: the Faculty of Humanities at the University of Pretoria.

## Authors contributions

SMN, LN-A and FZ conducted the preliminary database search for the protocol, conceptualised, drafted, edited, reviewed, sent out the draft protocol manuscript for constructive feedback, and approved the final manuscript.

## Ethics approval and consent to participate

Not applicable.

## Consent for publication

Not applicable.

## Availability of data material

The required data is included in the manuscript.

## Abbreviations

ACHPR: African Commission on Human and People’s Rights
HEI: Higher Education Institutions
ILGA: International Lesbian, Gay, Bisexual, Trans and Intersex Association
LIC: Low Income Countries
LMIC: Lower-Middle-Income Countries
MMAT: Mixed-Method Appraisal Tool
NGO: Non-governmental organisation
NPO: Non-profit organisation
P.I.Co: Population. Interest. Context
PRISMA: Preferred Reporting Items for Systematic and Meta Analyses
PTSD: Post-Traumatic Stress Disorder
PEPFAR: the United States Presidents Emergency Plan for Aids Relief
SSA: sub-Saharan Africa
SDG: Sustainable Development Goals
UN: United Nations
UNAIDS: the Joint United Nations Programme on HIV/AIDS
UNFPA: the United Nations Population Fund
UNESCO: the United Nations Educational, Scientific and Cultural Organization
UN-Habitat: United Nations Human Settlements Programme
WHO: World Health Organization

---

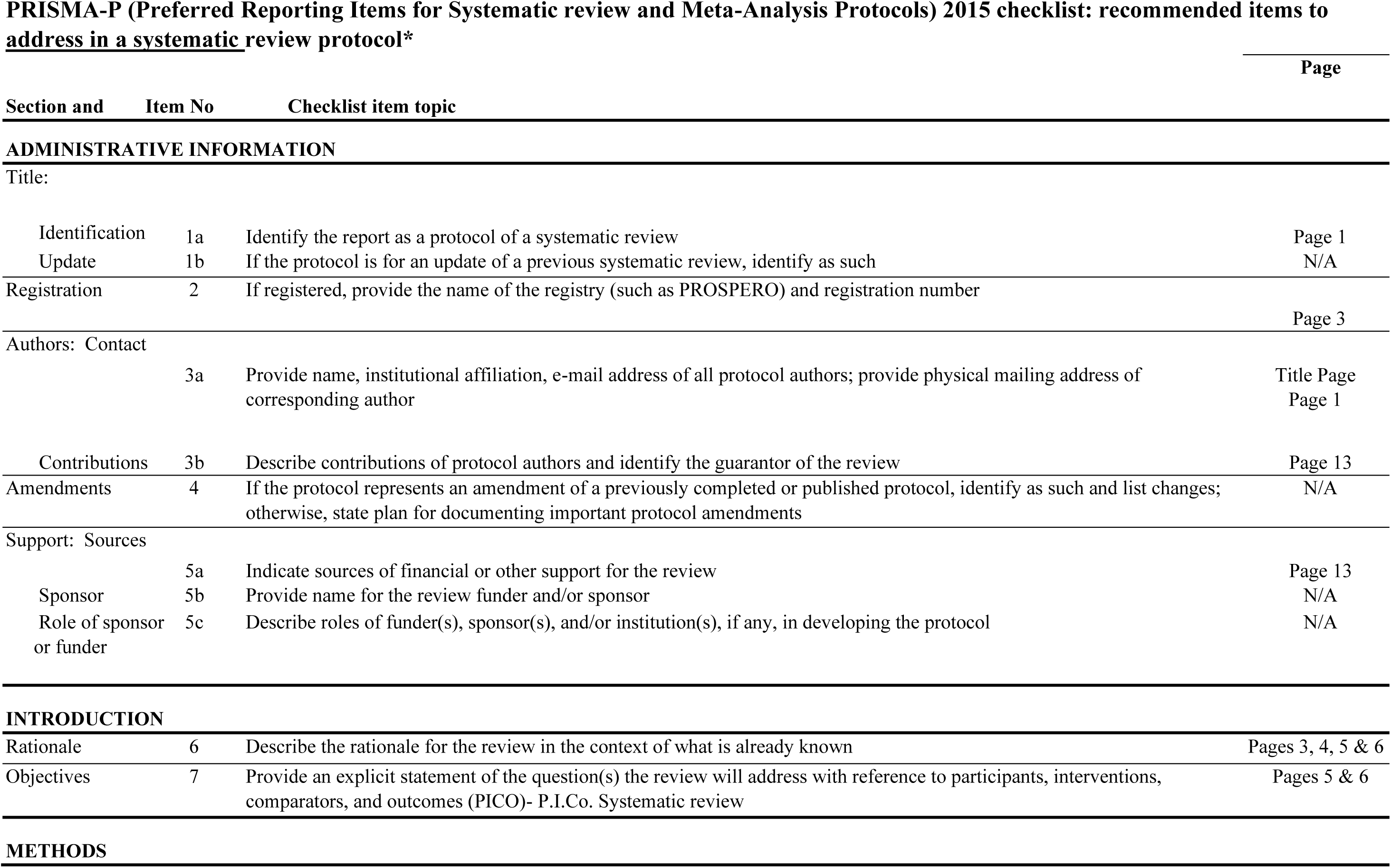
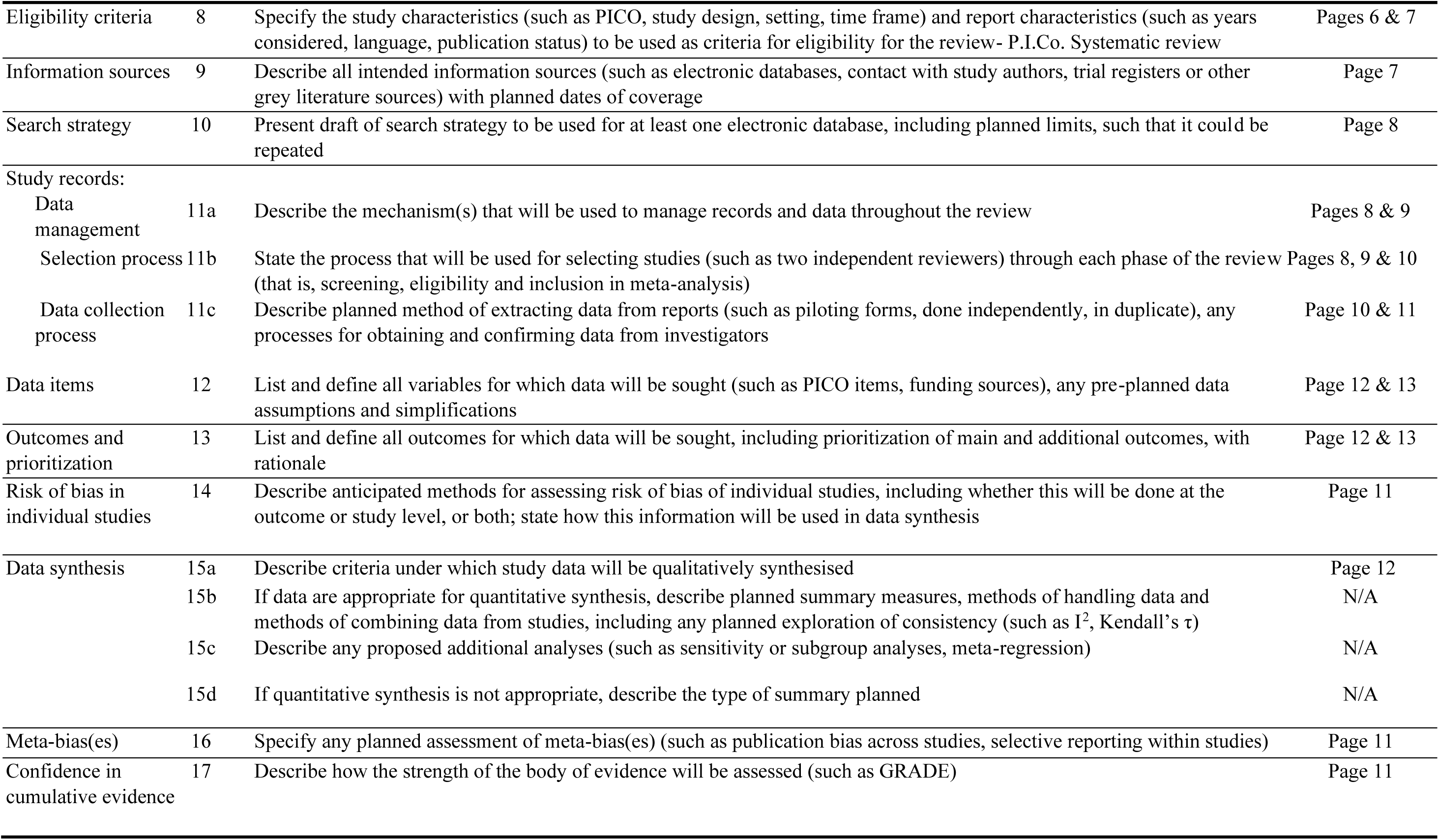
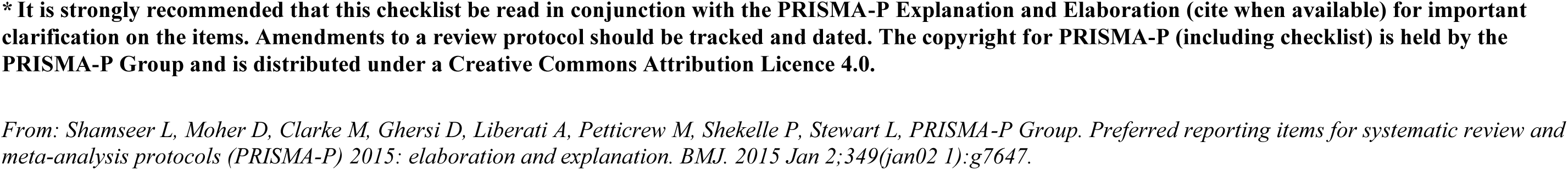

